# Statistical Analysis Plan for the manuscript reporting the primary outcome in the TESSA study

**DOI:** 10.1101/2025.01.03.24318812

**Authors:** Louise Groth Grunnet, Jonas Salling Quist, Kristine Færch, Martin Bæk Blond, Andre Kengne, Joel Dave, Julia Goedecke, Amy Mendham

## Abstract

This paper describes the statistical analyses plan (SAP) for the following study: Exploring the effects of time-restricted <u>e</u>ating on body weight and associated cardiometabolic outcomes in <u>S</u>outh <u>A</u>frican women living with HIV (**TESSA**): a protocol for a randomised controlled trial.

This document is a supplement to the TESSA study protocol and contains the statistical analysis plan (SAP) for the main paper of the trial in which the primary outcome will be reported. This document complies with the guidelines for the content of statistical analysis plans in clinical trials. The SAP does not cover the statistical analysis of data for outcomes to be included in secondary manuscripts.

## Section 2: Introduction

### 7 Background and Rationale (adapted from study protocol)

By 2045, sub-Saharan Africa is projected to see a 134% increase in type 2 diabetes (T2D) to 55 million cases, with South Africa among the highest prevalence regions (1). T2D is the leading cause of death in South African women, contributing to 7.2% of deaths (1.2). South Africa also bears the highest global HIV burden (7.7 million), disproportionately affecting women (3). Since 2019, dolutegravir (DTG) has been the WHO-recommended first-line HIV treatment. However, transitioning to DTG-based regimens has been linked to significant weight gain, raising concerns about its impact on health and the need for weight management interventions.

Lifestyle interventions effective in high-income countries are challenging to adapt to resource-limited settings like South Africa due to systemic and personal barriers such as food insecurity, poverty, and low access to quality healthcare. With 35% of South African households experiencing food insecurity in 2021, traditional dietary strategies are difficult to implement. Time-restricted eating (TRE), limiting eating to 8–10 hours daily, may offer a low-cost, feasible weight management approach. TRE has shown promising cardiometabolic benefits in developed countries (4,5), though data from resource-poor communities is limited.

This study, based on co-design research and pilot phases, investigates the impact of TRE on weight and cardiometabolic outcomes in South African women with overweight/obesity and HIV on DTG-based ART. It hypothesizes that an 8–10-hour TRE intervention can prevent weight gain and improve health outcomes in this high-risk population.

### 8 Objectives

The primary objective of the TESSA study is to investigate the effects of TRE on changes in body weight from baseline to 12 months in South African women living with overweight/obesity and HIV receiving DTG-based ART. Secondary objectives are to describe changes associated with the intervention for body composition, cardiometabolic risk and behaviour.

#### Hypotheses for the primary outcome

1. We hypothesize that TRE is superior to no intervention (the control group) in preventing weight gain from baseline to end-of-treatment (from baseline to 12 months). Superiority is claimed if:
  a. the 95% confidence interval for the estimated difference in change between the groups for total body weight, estimated using a baseline corrected linear mixed model (see Section 6.27), excludes 0 and the P-value is < 0.05; and
  b. the estimated difference in total body weight between the two groups is equal to/surpass the minimal important difference in favour of the TRE group.

## Section 3: Study Methods

### 9 Trial design

Single-centre parallel-group, randomized controlled, open-label, superiority trial. Allocation ratio 1:1 to either TRE + DTG treatment or control (no-intervention + DTG treatment).

### 10 Randomization

Participants will be randomized 1:1 using REDCap. An international collaborator not involved in data collection generated and uploaded the randomization list to the electronic data management system REDCap (8.10.18, Vanderbilt University, TN, USA).

### 11 Sample size

Sample size calculation was completed based on a power of 80%, a standardized mean difference of 0.5 between the two groups (medium effect size), α=0.05 requires a sample size of 128 (64 per group), but with an expected drop-out rate of 20%, we will include 152 individuals (76 in the TRE intervention group and 76 in the control group) in the RCT.

### 12 Framework

Superiority trial. See point 8

### 13 Statistical interim analyses and stopping guidance

No interim analyses are planned and no guidelines for terminating the trial early has been made.

### 14 Timing of final analyses

Results will be analyzed when the last participant completes the last visit.

### 15 Timing of outcome assessments

Body weight is measured at the baseline test and monthly until 12 months when the intervention ends. See item 25 for timing of secondary and descriptive/explorative outcome assessments.

## Section 4: Statistical Principles

### 16-18 Confidence intervals and P values

Two-sided P-values and 95% confidence intervals will be presented for comparisons (between and within groups). Likewise, two-sided 95% confidence intervals will be presented for estimated levels.

#### Primary outcome

Statistical significance will be claimed if the null hypothesis is rejected at the alpha level of 0.05 (two-sided), i.e. the P-value of the null hypothesis test is < 0.05. The direction and size of the estimated mean effect for total body weight (primary outcome), in addition to the 95% confidence intervals, will be required to support the tested hypothesis in order for the results to be declared in accordance with the hypothesis (see item 8).

#### Secondary outcomes

False detection rate (FDR) correction ad modem Benjamini and Hochberg (5) will be used to control for multiplicity; < 5% will be used as the threshold for FDR.

#### Descriptive/explorative outcome assessments

No correction for multiplicity is planned and non-hypothesis based tests are per definition of a descriptive/exploratory nature from which no definite inferences can be made.

### 19 Adherence and protocol deviations

#### Compliance

%-compliance = (number of days compliant / total days) * 100%. Compliant days are defined as:

- not exceeding the 10-hour eating duration by more than 1 hour daily (i.e., have an eating duration < 11 hours/day); and
- starting less than one hour before and ending less than one hour after the 10-hour self-selected eating window. Eating window delimited by the self-selected start and end of daily intake of food and beverages (except of water, black tea or coffee, with no sweeteners, sugar or milk).

Days for which a participant has not recorded eating window and/or duration will be regarded as non-compliant days. No compliance criteria for the participants in the control group have been defined.

#### Completers

Participants who participate in assessment of primary outcome at 12 months.

#### Lost to follow-up

Participants who do not participate in assessment of primary outcome at 12 months.

#### Per protocol

Completers in the TRE group are considered *per protocol* if their %-compliance is ≥80%. All completers in the control group are considered per protocol.

#### Protocol deviators

All completers in the TRE group, who are not considered per protocol. No participants in the control are considered protocol deviators.

#### Summary data/presentation of data

Adherence to the intervention in the TRE group will be presented as the summary data of percentage of days for which the participants are compliant. Additionally, the average eating duration (hours:minutes) will be presented. The average volume of time eating outside the self-selected eating window (hours:minutes) for the TRE group will be presented. Data related to measures of adherence will be presented for all completers and for per protocol completers in the TRE group. The distribution of the outcome will be visually inspected using QQ-plots and histograms and if Gaussian distributed, will be presented as means and standard deviations; if not the data will be presented as medians plus 25^th^ and 75^th^ percentiles.

### 20 Analysis populations

#### Intention-to-treat (ITT) analysis set

All participants will be analyzed as randomized.

#### Per Protocol (PP) analysis set

See item 19 for definition of per protocol.

## Section 5: Trial Population

### 21 Eligibility

Inclusion Criteria

- Women
- Age: ≥20to ≤45 years
- Body mass index ≥25 kg/m2
- Living with HIV and on DTG treatment for 1-24 months

Exclusion criteria

- Active or unstable medical conditions affecting body weight or glycaemic control
- Severe eating disorders
- Significant psychiatric illnesses
- Inflammatory bowel disease
- Malabsorption-related disorders
- Known active tuberculosis
- Helminth infection
- Hepatitis
- Alcohol/drug addiction
- Pregnant, breastfeeding or planning pregnancy within a year
- Postmenopausal
- Recent positive COVID-19 tests or current COVID-19 symptoms
- Over 5% weight loss during the last 6 months
- Use of medications such as glucocorticoids, hydrochlorothiazide and beta-blockers
- Concurrent involvement in another intervention study

### 22 Recruitment

The flow chart of the trial will comply with the CONSORT guidelines and will include the number of individuals who:

1. received oral information
2. were assessed for eligibility at screening; including the number eligible and the number ineligible
3. consented
4. participated in baseline tests
5. were randomized
6. attended each of the visits
7. completed per protocol at 3, 6, 9 and 12 months
8. were included in the primary analysis

Additionally, the attrition, and if possible, reasons for attrition will be supplied for each phase of the trial, and if applicable the number of per protocol completers will be given.

### 23 Withdrawal/follow-up

The level of consent and consent withdrawal will be tabulated (classified as “consent to continue follow-up and data collection”, “withdrawal of consent – no further follow-up or data collection”). If a randomized participant wishes to withdraw from the trial, they will be contacted to participate in a call which tries to find out their reason for withdrawing from the trial. Every reason for withdrawal is captured on REDCap. The number of participants lost-to-follow-up for each group during each phase of the trial will be reported in the CONSORT diagram (See item 23). Summary of baseline characteristics of completers and those lost to follow-up will be provided in the baseline table. Spaghetti plots will be used to visualize levels of the main outcome for completers and non-completers.

### 24 Baseline participant characteristics

The distribution of all continuous outcomes included in baseline characteristics will be visually inspected using QQ-plots and histograms; those with a Gaussian distribution will be presented as means and standard deviations and those with a non-Gaussian distribution will be presented as medians plus 25^th^ and 75^th^ percentiles, and number of observations will be presented for each outcome presented. Categorical data will be summarised by numbers and percentages. Tests of statistical significance will not be undertaken for baseline characteristics; rather the clinical importance of any imbalance will be noted.

The following variables will be included in the baseline participant characteristics table for all participants combined and stratified by randomization group:

- Number of women
- Age (years)
- Self-reported ethnicity (n and %)
- Current smoker, n (%)
- Education
  - Grade 7 and less (n and %)
  - Grade 8-12 (High School) (n and %)
  - Matric and above (n and %)
- Occupation
  - Employed (n and %)
  - Unemployed (n and %)
  - A student (n and %)
  - Informal (n and %)
  - Other (n and %)
- Grants
  - Disability (n and %)
  - Social support (n and %)
  - None (n and %)
- People living in household, n
- House type
  - House built formal/ brick house (n and %)
  - Informal shack / shelter / zozo (n and %)
- Medical History of disease and Medication use and family history of disease
- Weight (kg)
- BMI (kg/m^2^)
- Waist circumference(cm)
- Waist-hip ratio (unitless)
- Total fat mass (kg)
- Total fat free mass (kg)
- Body fat percentage (%)
- Average daily eating duration (hours:minutes)
- Total energy intake (KJ/day and/or kcal/day)
- Macronutrient intake
  - Carbohydrate (KJ/day and percentage of total energy intake (E%))
  - Fat (KJ/day and E%)
  - Protein (KJ/day and E%)
  - Alcohol (KJ/day and E%)
- Systolic blood pressure (mmHg)
- Diastolic blood pressure (mmHg)
- HbA1c (mmol/mol and %)
- Fasting glucose (mmol/L)
- Fasting insulin (pmol/L)
- Fasting total Cholesterol (mmol/L)
- Fasting HDL-Cholesterol (mmol/L)
- Fasting LDL-Cholesterol (mmol/L)
- Fasting Triglycerides (mmol/L)
- Pittsburgh Sleep Quality Index (PSQI)
  - Global score (0-21)
  - Sleep duration (hours/min)
- Quality of life (EQ5D)
- Physical activity (GPAQ)
- Household Food insecurity (Household Food insecurity Access Scale, HFIASA)
- Perceived appetite and hunger (VAS)
- Social Support for Diet and Exercise Behaviours

## Section 6: Analysis

### 25 Outcome definitions

#### Primary outcome

Total body weight (measured to the nearest 0.1 kg). The treatment effect will be given as the baseline corrected difference in kg between the groups at 12 months. The relative difference (%) will also be provided. The minimally important difference (MID) is defined as Cohen D at 0.5 and will be calculated as 0.5 *SD measured at baseline in all randomized individuals.

#### Secondary outcomes

The treatment effect will be given as the baseline corrected difference <u>between the groups at 12 months</u>. No minimal clinically relevant differences were defined for these outcomes.

- Total body fat mass (kg)
- Total fat free mass (kg)
- Total energy intake (KJ/day)
- HbA1c (mmol/mol and %)
- Fasting plasma glucose (mmol/L)
- Fasting concentrations of LDL, TG, HDL and TC
- Matsuda index - measure of insulin sensitivity
- Insulinogenic index (insulin 30-0/glucose 30-0) – estimate of insulin response
- Disposition index (Matsuda x insulinogenic index) – estimate of beta-cell function
- HOMAIR and HOMAB
- Systolic blood pressure (mmHg)
- Diastolic blood pressure (mmHg)
- Quality of life – EQ5D
- Self efficacy
- Sleep quality – PSQI
- Global score (0-21)
- Sleep duration (hours/min)
- Dietary intake
  - Total energy intake (KJ/day and/or kcal/day)
  - Carbohydrate (KJ/day and percentage of total energy intake (E%))
  - Fat (KJ/day and E%)
  - Protein (KJ/day and E%)
  - Alcohol (KJ/day and E%)
- Physical activity (GPAQ) (Minutes moderate to vigorous, light activity and sedentary)
- VAS (hunger, appetite, cravings, satiety)

#### Descriptive/explorative outcomes

Baseline corrected difference at <u>3, 6 and 9 months</u> for:

The above-mentioned outcomes and addition food insecurity, TRE compliance

Furthermore, a qualitative process evaluation will be performed with structured interviews at 3, 6, 9 and 12 months.

### 26 Analysis methods

Analyses of the primary outcome (except for the pre-planned per protocol) will be performed based on the intention-to-treat (ITT) principle.

Before further analysis and before unblinding, all variables will be inspected to detect outliers in order to uncover potential errors, such as registration errors. Data for all participants with a change from baseline of more than 5% of total body weight will checked to rule out registration error. In the case of evidence of a registration error, this will be corrected, otherwise the value will remain unchanged.

All continuous endpoints/outcomes covered by this SAP will as a rule be modelled using baseline corrected repeated measures regression (6) with the following fixed effects and interactions between fixed effects: Treatment, Visit (factorial)*Treatment. Data from baseline, 3, 6, 9 and 12 months will be included in the analysis. All participants will be placed in the control group at baseline in the statistical analysis. The models will be specified with a restricted maximum likelihood estimation method and a repeat on participant level (unstructured covariance structure). Model fit will be evaluated using graphical methods before estimating the treatment effects and if necessary, outcomes will be log-transformed. Estimated mean differences (CI95%) between groups, conditional means (CI95%), and within group changes (CI95%) will be extracted from the model. For log-transformed outcomes the results will be back-transformed and be presented as the ratio between estimated mean differences (CI95%), estimated conditional geometric means (CI95%) and relative changes within groups (CI95%), respectively. If distribution assumptions cannot be met by log-transformation, a generalized mixed linear model with an appropriate distribution will be applied instead of the repeated measures regression model. In case the distribution does not comply with the distributions available in the generalized mixed linear model a non-parametric test will be used to compare the change scores for the given outcome.

Between group differences will be null-hypothesis tested and presented with P-values.

The following two supplementary analyses are planned for the primary outcome:

1. An analysis similar to the one described above but including the per-protocol analysis population.
2. A sensitivity analysis, including the ITT analysis population, assessing the impact of missing data for the primary outcome. This analysis will only be performed in the case of a loss-to-follow-up rate > 20%, i.e., more than 20% of all participants are missing at 12 months (see also item 27).

The analysis of the primary and secondary outcomes will be performed blinded to group allocation by a researcher that has not been involved in the execution of the trial.

### 27 Missing data

The number/frequency of missing values for the primary outcome in each group at each time point will be provided. In the primary analysis, missing data are handled implicitly by maximum likelihood estimation in the linear mixed model and missing data will be assumed to be missing at random. This is in the model specified in point 27 equivalent to making multiple imputations for each treatment group separately and estimates the treatment effect that would have been found had all subject completed their assigned treatment (efficacy estimate) under the missing at random assumption. In the case that >20% of the participants are lost-to-follow-up and/or >20% in one of the groups, we will perform a supplementary analysis to challenge the assumptions of “missing at random” in the primary analysis. We will assume that the development in body weight among the participants from TRE that are lost-to-follow-up will resemble the development in the control group rather than the development in the group to which they were originally randomized. Participants from the TRE group with missing values at V3 and/or V4 will be pooled with the control group and using a Markov chain Monte Carlo method all missing values will be imputed to create 1000 new datasets, assuming a multivariate normal distribution for the data. The dataset used for the imputation will include all previous measurements for weight (excluding that from the screening visit), age and sex. Subsequently, the imputed datasets will be analyzed using a mixed linear model including the same variables included in the main analysis and averaged estimates will be calculated. If the main outcome has been transformed to fit the statistical model used for the main analysis this transformation will be applied to the outcome prior to the imputation procedure.

### 28. Additional analyses

Not relevant

### 29 Harms

Data on harms are not systematically collected and will not be reported.

### 30 Statistical software

R version 3.6.0 or newer version (The R Foundation for Statistical Computing, www.R-project.org) and SAS version 9.4 or newer version (SAS Institute, Cary, NC, USA).

## Data Availability

All data produced in the present study are available upon reasonable request to the authors

